# Genetic Predictors of Blood Pressure Traits are Associated with Preeclampsia

**DOI:** 10.1101/2023.02.09.23285734

**Authors:** Elizabeth A. Jasper, Jacklyn N. Hellwege, Joseph H. Breeyear, Brenda Xiao, Gail P. Jarvik, Ian B. Stanaway, Kathleen A. Leppig, Geetha Chittoor, M. Geoffrey Hayes, Ozan Dikilitas, Iftikhar J. Kullo, Ingrid A. Holm, Shefali Setia Verma, Todd L. Edwards, Digna R. Velez Edwards

**Affiliations:** Department of Obstetrics and Gynecology, Vanderbilt University Medical Center, Nashville, TN, U.S.; Department of Biomedical Informatics, Vanderbilt University Medical Center, Nashville, TN, U.S.; Vanderbilt Genetics Institute, Vanderbilt University, Nashville, TN, U.S.; Vanderbilt Epidemiology Center, Vanderbilt University, Nashville, TN, U.S.; Institute for Medicine and Public Health, Vanderbilt University Medical Center, Nashville, TN, U.S.; Division of Genetic Medicine, Department of Medicine, Vanderbilt University Medical Center; Department of Genetics, University of Pennsylvania, Philadelphia, PA, U.S.; Departments of Medicine (Medical Genetics) and Genome Sciences, University of Washington Medical Center, Seattle, WA, U.S.; Department of Medicine, Division of Nephrology and Harborview Medical Center Kidney Research Institute, University of Washington Medical Center, Seattle WA, U.S.; Genetic Services, Kaiser Permanente of Washington, Seattle, WA, U.S.; Department of Population Health Sciences, Geisinger, Danville, PA, U.S.; Division of Endocrinology, Metabolism, and Molecular Medicine, Department of Medicine, Northwestern University Feinberg School of Medicine, Chicago, IL, U.S.; Center for Genetic Medicine, Northwestern University Feinberg School of Medicine, Chicago, IL, U.S.; Department of Anthropology, Northwestern University, Evanston, IL, U.S.; Departments of Internal Medicine, Cardiovascular Medicine, Mayo Clinician-Investigator Training Program, Mayo Clinic, Rochester, MN, U.S.; Department of Cardiovascular Medicine, Mayo Clinic, Rochester, Minnesota, US; Division of Genetics and Genomics, Boston Children’s Hospital, Boston, MA, U.S.; Department of Pediatrics, Harvard Medical School, Boston, MA, U.S.; Department of Pathology and Laboratory Medicine, University of Pennsylvania, Philadelphia, PA, U.S; Division of Epidemiology, Department of Medicine, Vanderbilt University Medical Center, Nashville, TN, U.S.

**Keywords:** Polygenic risk, genetic epidemiology, pregnancy health, preeclampsia, blood pressure

## Abstract

**Background:** Preeclampsia, a pregnancy complication characterized by hypertension after 20 gestational weeks, is a major cause of maternal and neonatal morbidity and mortality. The mechanisms leading to preeclampsia are unclear; however, there is evidence that preeclampsia is highly heritable. We evaluated the association of polygenic risk scores (PRS) for blood pressure traits and preeclampsia to assess whether there is shared genetic architecture.

**Methods:** Participants were obtained from Vanderbilt University’s BioVU, the Electronic Medical Records and Genomics network, and the Penn Medicine Biobank. Non-Hispanic Black and White females of reproductive age with indications of pregnancy and genotype information were included. Preeclampsia was defined by ICD codes. Summary statistics for diastolic blood pressure (DBP), systolic blood pressure (SBP), and pulse pressure (PP) PRS were obtained from Giri et al 2019. Associations between preeclampsia and each PRS were evaluated separately by race and study population before evidence was meta-analyzed. Prediction models were developed and evaluated using 10-fold cross validation.

**Results:** In the 3,504 Black and 5,009 White individuals included, the rate of preeclampsia was 15.49%. The DBP and SBP PRSs were associated with preeclampsia in Whites but not Blacks. The PP PRS was significantly associated with preeclampsia in Blacks and Whites. In trans-ancestry meta-analysis, all PRSs were associated with preeclampsia (OR_DBP_=1.10, 95% CI=1.02-1.17, *p*=7.68×10^−3^; OR_SBP_=1.16, 95% CI=1.09-1.23, *p*=2.23×10^−6^; OR_PP_=1.14, 95% CI=1.07-1.27, *p*=9.86×10^−5^). However, addition of PRSs to clinical prediction models did not improve predictive performance.

**Conclusions:** Genetic factors contributing to blood pressure regulation in the general population also predispose to preeclampsia.

## Introduction

Preeclampsia, a pregnancy complication characterized by hypertension after 20 gestational weeks, is a major cause of maternal and neonatal morbidity and mortality. Globally, preeclampsia is estimated to complicate 2.0% to 8.0% of all pregnancies^1^. In the United States (U.S.), the prevalence of preeclampsia increased by 25% between 1987 and 2004^2^. The risk of preeclampsia has also increased over the years, with women giving birth in 2003 at a 6.7-fold increase risk of preeclampsia compared to women who gave birth in 1980^3^. In addition to the rising rates and risk of preeclampsia, the condition is costly. In the U.S. in 2012, one study estimated the cost of preeclampsia in the year following delivery to be $2.18 billion dollars^4^. In addition to its immediate economic cost, the pregnancy complication has massive effects on the health of the mother and infant. Risk of stillbirth, neonatal death, fetal growth restriction, lower birth weights, preterm birth, and perinatal mortality is higher for women with preeclampsia^5,6^. There is growing evidence that preeclampsia is not just an issue of pregnancy, rather it effects the long-term health of mothers and infants. Preeclamptic women are later at increased risk of developing and dying from cardiovascular disease while children exposed to preeclampsia have higher blood pressures, body mass index (BMI), and risk of hypertension^5,7^.

Pre-existing maternal conditions, pregnancy-specific characteristics, environmental factors, and family history have been identified as risk factors for preeclampsia. Increasing age of mothers, higher BMI, pre-existing hypertension, and diabetes all place women at increased risk of preeclampsia during pregnancy^8-11^. Having preeclampsia in a prior pregnancy and/or a family history of preeclampsia are significant risk factors for preeclampsia, with odds of preeclampsia in the current pregnancy being 2.9 (95% confidence interval (CI): 1.70-4.93) times greater in individuals who previously had preeclampsia and 7.19 (95% CI: 5.85-8.83) times more likely in those with family who had preeclampsia compared to/than those without personal or family history^9-11^. Heritability studies of preeclampsia present strong support for a genetic basis for preeclampsia with twin studies estimating heritability for preeclampsia to be approximately 55%^12^. Several small genome wide association studies (GWAS) have been performed, with less than 50 associated polymorphisms identified to date^13-15^. However, identified risk factors and polymorphisms have yet to fully explain the exact biologic mechanism leading to preeclampsia. There is some evidence supporting that genetic predisposition to hypertension may be associated with preeclampsia in European ancestry and Asian women^14^. Several polymorphisms associated with preeclampsia have also been previously associated with blood pressure traits, though the shared genetic risk has yet to be fully evaluated^14,15^.

As preeclampsia is a blood pressure disorder and puts individuals at greater risk of hypertension later in life, we investigated the potential shared genetic architecture of other blood pressure traits and preeclampsia in EHR-identified Black and White women. To increase the understanding of preeclampsia etiology and identify the potential shared genetic architecture with other blood pressure traits, we evaluated the association between polygenic risk scores (PRS) for three blood pressure traits—diastolic blood pressure (DBP), systolic blood pressure (SBP), and pulse pressure (PP)—and preeclampsia in several cohorts of Black and White reproductive-aged females with documented pregnancies. We also investigated the utility of blood pressure PRSs for the prediction of preeclampsia, determining whether the addition of genetic components improve the performance of prediction models composed of clinical risk factors.

## Methods

### Data Availability

The data that support the findings of this study are available from the corresponding author upon reasonable request. Data from the Penn Medicine Biobank is available from Dr. Verma upon reasonable request.

### Study Populations

Our study populations were obtained from three sources: BioVU, Vanderbilt University Medical Center’s biorepository linking DNA samples to deidentified electronic health records (EHR), the Electronic Medical Record and Genomics Network (eMERGE), a national network combining DNA biorepositories with EHR systems, and Penn Medicine Biobank (PMBB) at the University of Pennsylvania^16-18^. This study was approved by Vanderbilt University Medical Center Institutional Review Board. Individuals were eligible for inclusion in the study if they were women who had documented pregnancies or deliveries, were of reproductive age at last known EHR entry (18 to 45 years old), identified in the EHR as non-Hispanic Black or non-Hispanic White and had genotype information available. Identification of individuals who are or were pregnant and/or had deliveries was accomplished using International Classification of Disease, 9^th^ and 10^th^ revisions (ICD-9/ICD-10) codes. Presence of any of the following codes indicated potential eligibility for inclusion in the study: 631, 633, 634.3, 635 through 679, 760-779, 796.5, V22-V24, V27, V28, V72.42, V82.4, V89, V91 (ICD-9), O09-O16, O20-O26, O28-O36, O40-O48, O60-O77, O80, O82, O85-O92, O94, O98, O99, O9A, Z03.7, Z32.01, Z33, Z34, Z36, Z37, Z39, or Z3A (ICD-10). Preeclampsia cases were be defined using the presence of any one of codes 642.4, 642.5, 642.6, 642.7 (ICD-9 codes), O11, O14, and O15 (ICD-10).

### Blood Pressure Traits Polygenic Risk Scores (PRS) Development

Summary statistics for genetic associations with DBP, SBP, and PP were obtained from a previously published trans-ancestry genome-wide association study (n_max_ = 760,226 subjects)^19^. The SBP and DBP PRSs were validated in BioVU (n_max_ = 37,132, DBP PRS *p* = 7.60×10^−113^, SBP PRS *p* = 4.18 ×10^−132^) for association with blood pressure measurements, adjusting for age, sex, BMI, and the top ten principal components of ancestry^20^. The BioVU genetic data was pruned for linkage disequilibrium at an r^2^ threshold of 0.1 with a maximum distance of 250 kilobases from associated single nucleotide polymorphisms (SNPs) (p < 1 ×10^−5^) in the blood pressure summary statistics. Weighted scores were calculated in PLINK using p-value thresholding^21^. The DBP, SBP, and PP scores contained 4,194, 4,294, and 3,345 SNPS respectively (Supplemental Tables 1-3)^20^. PRS were transformed by multiplying scores by the standard deviation of the PRS in the original population from which they were optimized.

**Table 1:**
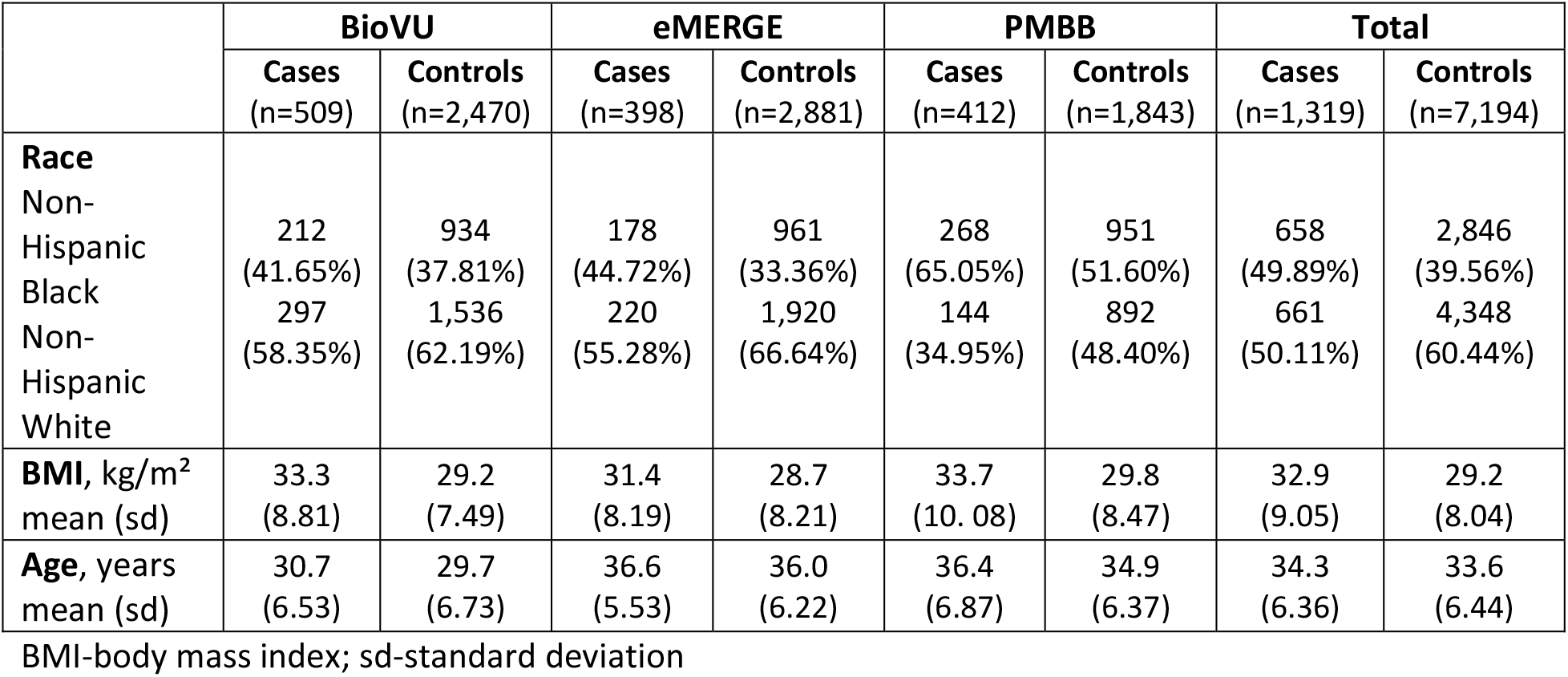
Population characteristics

### Blood Pressure Trait PRSs-Preeclampsia Association Analyses

Logistic regression was used to evaluate the association between each of the three blood pressure traits and preeclampsia separately in the three cohorts (BioVU, eMERGE, and PMBB) stratified by race (Black and White). Analyses were adjusted for age, age squared, and the top ten principal components. Effect estimates and standard errors from the three populations were pooled in race-stratified and trans-ancestry fixed-effects meta-analyses using an inverse-variance approach in RevMan, version 5.4^22^. Associations were considered statistically significant at *p*-values less than 0.05. Odds ratios (OR) and 95% confidence intervals (CI) are reported for each model as the increase in risk of preeclampsia per one standard deviation increase in of each blood pressure polygenic risk score. Analyses were completed using R version 4.2.2^23^.

### Predictive Modeling

To evaluate the performance of blood pressure PRS for the prediction of preeclampsia, we developed prediction models using 10-fold cross validation. Joint prediction models composed of all three PRSs (DBP, SBP, and PP) were developed. Clinical models, composed of two major preeclampsia risk factors (BMI and age at last EHR entry), were constructed for comparison. A full model, which added the three PRSs to the clinical model, was used to investigate whether PRS improved prediction of preeclampsia beyond traditional risk factors. Models that included PRSs also adjusted for the top ten principal components.

Models were developed in the cohort from eMERGE and tested in BioVU. Synthetic minority over-sampling techniques (SMOTE) were used to attenuate the effects of class imbalance (number of cases and controls) due to the rarity of the outcome during model training^24^. Only those with complete covariate information were used in prediction modeling. All models were created and evaluated separately for each EHR-identified race (non-Hispanic Blacks and Whites) with subsequent development of trans-ancestry models. Model performance was assessed through discrimination and calibration using the area under the receiver operating characteristics curve (AUC) and the Brier Score. All prediction modeling was performed in R^23^.

## Results

### Population Characteristics

In total, 8,513 individuals were included in the study; 2,979 individuals from BioVU, 3,279 from the eMERGE cohort, and 2,255 from the PMBB cohort (Table 1). In the overall cohort, 41.16% were Black (38.47% in BioVU, 34.74% in eMERGE, 54.06% in PMBB cohort) and 58.84% were White (61.53% in BioVU, 65.26% in eMERGE, and 45.94% in PMBB). The overall rate of preeclampsia was 15.49%. Higher percentages of preeclampsia were found in Black individuals compared to White individuals (18.78% vs 13.20%). Within cohorts and across the entire population, cases had higher average BMIs and ages compared to controls.

### Blood Pressure Trait PRSs-Preeclampsia Associations

In Black individuals, PRSs for DBP and SBP were not significantly associated with preeclampsia in the meta-analyses (OR_DBP_ = 0.99, 95% CI = 0.88-1.12, *p*-value = 0.90; OR_SBP_ = 1.10, 95% CI = 0.99-1.23, *p*-value = 0.09) (Table 2). The PRS for PP was significantly associated with preeclampsia in Black individuals, with every one standard deviation increase in the PP PRS, the odds of preeclampsia is associated with a 1.16-fold increase (OR_PP_ = 1.16, 95% CI = 1.03-1.31, *p*-value = 0.01). In White individuals, DBP, SBP, and PP PRSs were all significantly associated with preeclampsia (OR_DBP_ = 1.14, 95% CI = 1.07-1.23, *p*-value = 2.25×10 ^-4^; OR_SBP_ = 1.18, 95% CI = 1.10-1.27, *p*-value = 4.42×10^−6^; OR_PP_ = 1.14, 95% CI = 1.05-1.23, *p*-value = 1.77×10^−3^). In the trans-ancestry meta-analysis, all three PRSs were highly significant (Table 3; Figures 1-3).

**Table 2:** Association between blood pressure PRSs and preeclampsia by self-reported race

|  | Black |  | White |  |
| --- | --- | --- | --- | --- |
|  | OR<br>(95% CI) | P-value | OR<br>(95% CI) | P-value |
| **DBP** | 0.99<br>(0.88, 1.12) | 0.89 | 1.14<br>(1.05, 1.23) | **1.21x10<sup>-3</sup>** |
| **SBP** | 1.10<br>(0.98, 1.22) | 0.11 | 1.18<br>(1.10, 1.27) | **4.24x10<sup>-6</sup>** |
| **PP** | 1.16<br>(1.03, 1.30) | **0.01** | 1.13<br>(1.05, 1.23) | **2.34x10<sup>-3</sup>** |

OR-odds ratio; CI-confidence interval; DBP-diastolic blood pressure; SBP-systolic blood pressure; PP-pulse pressure

**Table 3:** Association between blood pressure PRSs and preeclampsia (trans-ancestry)

|  | **OR (95% CI)** | **P-value** |
| --- | --- | --- |
| **DBP** | 1.10<br>(1.02, 1.17) | **$7.68 \times 10^{-3}$** |
| **SBP** | 1.16<br>(1.09, 1.23) | **$2.23 \times 10^{-6}$** |
| **PP** | 1.14<br>(1.07, 1.27) | **$9.86 \times 10^{-5}$** |

OR-odds ratio; CI-confidence interval; DBP-diastolic blood pressure; SBP-systolic blood pressure; PP-pulse pressure

**Figure 1.**
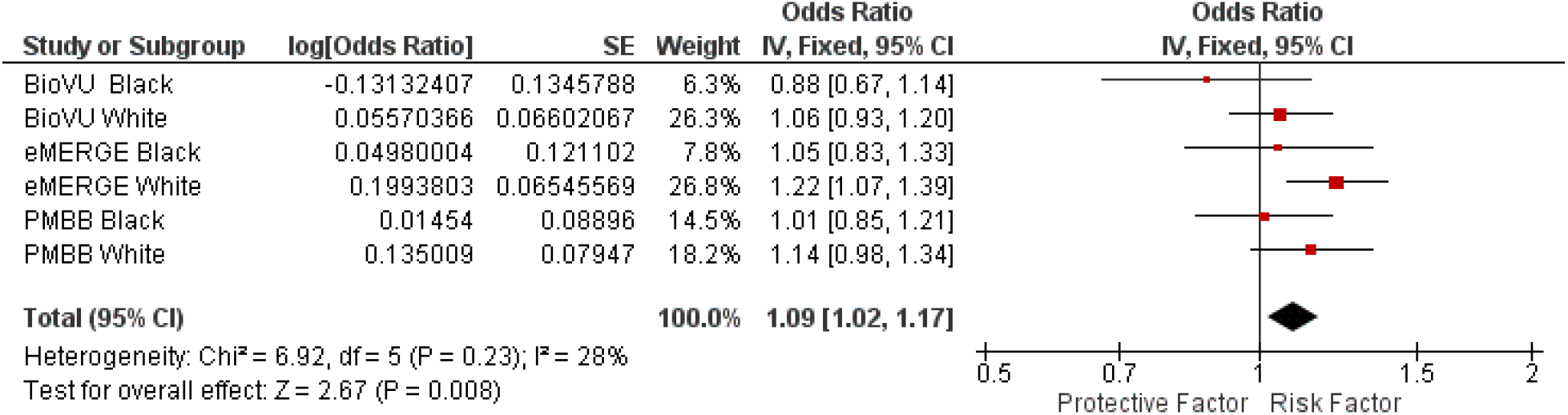
Trans-ancestry Meta-Analysis of the Association of Diastolic Blood Pressure Polygenic Risk Score and Preeclampsia

**Figure 2.**
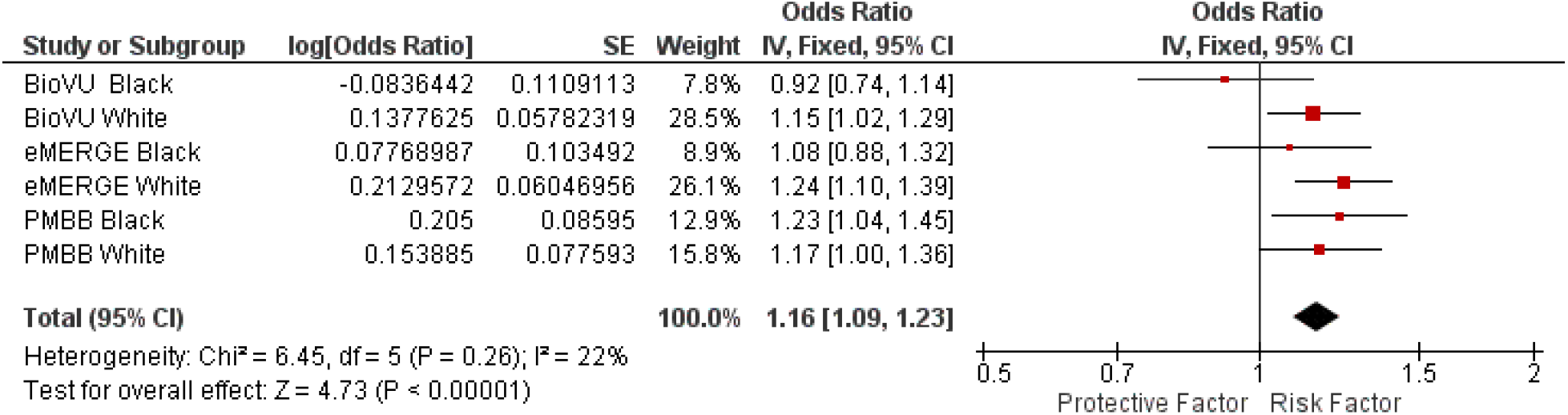
Trans-ancestry Meta-Analysis of the Association of Systolic Blood Pressure Polygenic Risk Score and Preeclampsia

**Figure 3.**
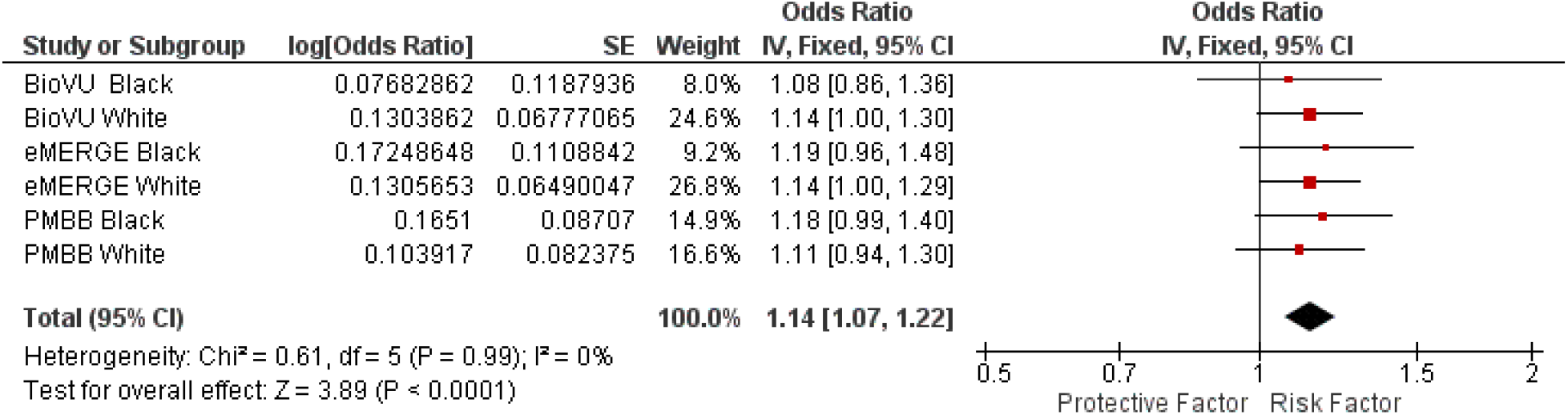
Trans-ancestry Meta-Analysis of the Association of Pulse Pressure Polygenic Risk Score and Preeclampsia

### Prediction Models

Models composed of solely of PRSs and 10 principal components had extremely poor discriminatory abilities, with AUCs ranging from 0.51 to 0.53 (Table 4). Clinical models had slightly better discrimination (AUCs: 0.59-0.67). The addition of the PRSs to the clinical models decreased the predictive ability of the models. Brier scores for PRS, clinical, and full models demonstrate low accuracy in the testing population. In general, lower AUCs and higher Brier scores were seen in PRS models compared to models composed of clinical risk factors and in EHR-identified non-Hispanic Black populations compared to White or trans-ancestry populations. Overall, inclusion of PRSs in a predictive model composed of significant clinical risk factors did not provide an improvement in predictive performance. Calibration and discrimination of the predictive models indicated poor fits in the testing population (Table 4).

**Table 4:** Prediction Models in testing (BioVU) population.

|  | Clinical model |  | PRS Model |  | Full Model |  |
| --- | --- | --- | --- | --- | --- | --- |
|  | AUC (95% CI) | Brier Score | AUC (95% CI) | Brier Score | AUC (95% CI) | Brier Score |
| **Black** | 0.5850<br>(0.5423-0.6277) | 0.3232 | 0.5181<br>(0.4742-0.5619) | 0.6409 | 0.5307<br>(0.4869-0.5744) | 0.6789 |
| **White** | 0.6732<br>(0.6368-0.7096) | 0.3034 | 0.5272<br>(0.4902-0.5641) | 0.2380 | 0.6590<br>(0.6232-0.6948) | 0.5234 |
| **Transethnic** | 0.6441<br>(0.6165-0.6717) | 0.2860 | 0.5091<br>(0.4803-0.5379) | 0.3036 | 0.5722<br>(0.5428-0.6017) | 0.3652 |

## Discussion

We evaluated the potential overlap between genetic risk factors for blood pressure traits and preeclampsia. Utilizing PRSs for DBP, SBP, and PP, we found significant associations between polygenic risk for these blood pressure traits and preeclampsia. These findings support shared genetic architecture across preeclampsia risk and blood pressure traits. Overall, these findings suggest genetic factors contributing to blood pressure regulation in the general population also predispose to preeclampsia.

However, polygenic risk scores for blood pressure traits on their own cannot fully predict preeclampsia status, as risk scores created using variants associated with these blood pressure traits had poor predictive ability. Furthermore, the addition of PRSs to a risk score composed of known clinical risk factors for preeclampsia did not result in a significant improvement in the prediction of preeclampsia.

Pre-pregnancy hypertension is a major risk factor for preeclampsia^8^. The development of preeclampsia also increases the risk of persistent hypertension outside of pregnancy^7^. This overlapping risk is likely due in part to shared underlying genetic mechanisms. Several preeclampsia GWAS support this idea, finding polymorphisms previously associated with hypertension are also associated with preeclampsia^14,15^. A recent large-scale meta-analysis of European and Central Asian women further reinforced the concept that genetic predisposition to hypertension contributes to the risk of hypertensive disorders of pregnancy^14^. While an association between a hypertension PRS and hypertensive disorders in pregnancy were observed, the effect of was stronger for gestational hypertension compared to preeclampsia, suggesting that, while there is overlap in genetic susceptibility, additional factors are involved in preeclampsia.

Variants associated with blood pressure traits have also been identified in previous GWAS for preeclampsia. The most recent large-scale study discovered two known blood pressure loci (in *FTO* and *ZNF831*) were significantly associated with preeclampsia^14^. Variants in several other blood pressure loci (*FGF5, LINC02356, MECOM, SH2B3, WLS, WNT3A*) in maternal genomes and a single variant in the *ITPR2* gene in fetal genomes showed suggestive associations with preeclampsia. A GWAS from the Hyperglycemia and Adverse Pregnancy Outcome study observed polymorphisms in *ADRA1D, RUNX1, ZNF295-AS1*, and *MUC21*/*MUC22*, which have previously been associated with diastolic, systolic, and/or pulse pressure, were also related to preeclampsia status. However, no SNP reached Bonferroni-corrected significance, likely due to the small sample size and subsequent limited power^15^.

It is unclear whether blood pressure complications and preeclampsia are comorbid conditions or if complications lead to preeclampsia or vice versa. Maternal cardiovascular systems undergo significant changes during pregnancy^25,26^. Individuals who experience preeclampsia may undergo further physiological changes. For example, previous research has noted significant increases in arterial stiffness in those with preeclampsia compared to individuals with normotensive pregnancy or those with gestational hypertension^27^. Our study provides further evidence of the role of arterial stiffness in preeclampsia, given that the PRS for PP, a marker of arterial stiffness, was strongly associated with preeclampsia in both Blacks and Whites. Additional pathways involved in preeclampsia could also be uniquely triggered by pregnancy-related changes and the fetal genome. Blood pressure regulation through genetic mechanisms may also be perturbed during pregnancy, with interactions resulting in those with increased genetic risk becoming more likely to develop preeclampsia. Additional research is needed to disentangle the causal origins of shared risk factors between blood pressure traits, complications, and preeclampsia and to identify variants that uniquely contribute to preeclampsia risk. Our cohort is also significantly younger than most cohorts used in blood pressure research. Age-related changes in blood pressure regulation have also been reported previously and could account for some of the variation seen between genetic risk factors for blood pressure traits and preeclampsia^28^.

This study builds upon previous research by investigating the traits underlying hypertension and further disentangling how genetic risk factors for these traits impact preeclampsia risk. Our results, which support the conclusion that genetic risk factors for blood pressure have a small contribution to the risk of preeclampsia, have several key strengths. First, several EHR-linked biorepositories were pooled in this analysis, increasing the sample size, power, and generalizability of results. The study cohort was also restricted to females with a pregnancy, ensuring all individuals possessed the appropriate window during which the outcome could have occurred. Previous research has been largely limited to Asian and White women or those of predominantly European ancestry. Here, we investigate preeclampsia genetic risk factors in EHR-identified Black individuals, a population known to be at increased risk of preeclampsia^11^.

Our study found small differences in the associations of blood pressure PRSs and preeclampsia between the EHR-identified races. These results should be interpreted carefully, especially in the non-Hispanic Black group which had small sample sizes and displayed a larger degree of uncertainty in effect estimates. The PRS were created using summary statistics from a trans-ancestry GWAS and the DBP and SBP PRS were subsequently validated on a trans-ancestry population. However, both populations were predominately those of European ancestry or non-Hispanic White individuals. The differences in the ancestry of the populations used to discover significant loci, those used to validate the PRS, and our study cohort could have biased the results and contributed to the lack of significance in the non-Hispanic Black populations. It is also notable that the PRS performed better in White compared to Black populations generally^20^, which may also impair the quality of their prediction of preeclampsia.

Prediction models composed of the three PRS had extremely poor predictive ability. Adding the PRS to the clinical model resulted in poorer performance than the clinical model alone. This could be due to a portion of individuals with higher genetic burden being classified as controls, as timing of hypertension development and diagnoses relative to pregnancy cannot be fully determined without prospective blood pressure measurement. Results from prediction modeling should also be considered cautiously as clinical and environmental risk factors, including as family history, were incomplete in these datasets. Additionally, many individuals in the study cohort were excluding during predictive model development and testing due to missing BMI. The addition of other known risk factors in development and testing of prediction models in datasets with more complete BMI information may increase their predictive ability and clinical value.

While this study further endorses the connection genetic risk factors for blood pressure and preeclampsia, results suggests that many variants, each with small effect sizes, likely affect genetic predisposition to preeclampsia. Though many of these variants may mainly impact preeclampsia through regulation of blood pressure, other molecular pathways are likely involved. Future research should continue to develop and validate genome informed clinical models for preeclampsia across global populations and conduct additional analyses of preeclampsia shared genetic architecture with other conditions. These analyses will expand our understanding of preeclampsia etiology and potentially inform future treatments by providing evidence for repurposing treatments for conditions that share a biological basis with preeclampsia.

## Acknowledgments

Drs. Elizabeth A. Jasper and Jacklyn N. Hellwege are supported by the NIH Building Interdisciplinary Research Career’s in Women’s Health career development program (K12HD043483 PIs: K.E. Hartmann, A.S. Major, and D.R. Velez Edwards). Dr. Jasper was also supported by a National Human Genome Research Institute award for the Vanderbilt Genomic Medicine Training Program (T32HG008341 PIs: J.F. Peterson, N. Cox, and D.M. Roden). Joseph H. Breeyear was supported by Vanderbilt University Medical Center’s Clinical and Translational Award training program (TL1-TR002244 PI: K.E. Hartmann). Iftikhar J. Kullo is funded by grants HG06379 and HG11710 from the National Human Genome Research Institute and K24HL137010 from the National Heart Lung and Blood Institute.

## BioVU

The dataset(s) used for the analyses described were obtained from Vanderbilt University Medical Center’s BioVU which is supported by numerous sources: institutional funding, private agencies, and federal grants. These include the NIH funded Shared Instrumentation Grant S10RR025141; and CTSA grants UL1TR002243, UL1TR000445, and UL1RR024975. Genomic data are also supported by investigator-led projects that include U01HG004798, R01NS032830, RC2GM092618, P50GM115305, U01HG006378, U19HL065962, R01HD074711; and additional funding sources listed at https://victr.vumc.org/biovu-funding/.

## eMERGE Network (Phase III)

This phase of the eMERGE Network was initiated and funded by the NHGRI through the following grants: U01HG008657 (Group Health Cooperative/University of Washington); U01HG008685 (Brigham and Women’s Hospital); U01HG008672 (Vanderbilt University Medical Center); U01HG008666 (Cincinnati Children’s Hospital Medical Center); U01HG006379 (Mayo Clinic); U01HG008679 (Geisinger Clinic); U01HG008680 (Columbia University Health Sciences); U01HG008684 (Children’s Hospital of Philadelphia); U01HG008673 (Northwestern University); U01HG008701 (Vanderbilt University Medical Center serving as the Coordinating Center); U01HG008676 (Partners Healthcare/Broad Institute); U01HG008664 (Baylor College of Medicine); and U54MD007593 (Meharry Medical College).

## eMERGE Network (Phase IV)

This phase of the eMERGE Network was initiated and funded by the NHGRI through the following grants: U01HG011172 (Cincinnati Children’s Hospital Medical Center); U01HG011175 (Children’s Hospital of Philadelphia); U01HG008680 (Columbia University); U01HG011176 (Icahn School of Medicine at Mount Sinai); U01HG008685 (Mass General Brigham); U01HG006379 (Mayo Clinic); U01HG011169 (Northwestern University); U01HG011167 (University of Alabama at Birmingham); U01HG008657 (University of Washington Medical Center); U01HG011181 (Vanderbilt University Medical Center); U01HG011166 (Vanderbilt University Medical Center serving as the Coordinating Center).

